# Circulating microvesicles as novel biomarkers for pulmonary arterial hypertension in patients with systemic lupus erythematosus

**DOI:** 10.1101/2024.03.10.24304030

**Authors:** Zhe Ding, Fumin Qi, Li Liu, Na Zhang, Xing Lyu, Wenwen Sun, Jun Du, Haoming Song, Hou Hou, Ying Guo, Xiaomei Wang, Ming-Lin Liu, Wei Wei

## Abstract

Pulmonary arterial hypertension (PAH) is a serious complication of systemic lupus erythematosus (SLE) with increased mortality. A prothrombotic state may contribute to pathogenesis of SLE-PAH. Microvesicles (MVs) are known to be associated with thrombosis. Here, we investigated circulating MVs and their associations with SLE-PAH. Eighteen SLE-PAH patients, 36 SLE-non-PAH patients, and 36 healthy controls (HCs) were enrolled. Flow cytometry was used to analyze circulating MVs from leukocytes (LMVs), red blood cells (RMVs), platelets (PMVs), endothelial cells (EMVs), and Annexin V^+^ MVs with phosphatidylserine (PS) exposure. Plasma levels of all MV subgroups were elevated in SLE patients with or without PAH compared to HCs. Furthermore, plasma Annexin V^+^ MVs, LMVs, PMVs, RMVs, EMVs, and Annexin V^+^ RMVs were significantly elevated in SLE-PAH patients compared to SLE-non-PAH patients. Additionally, PAH patients with moderate/high SLE showed a significant increase in LMVs, PMVs, RMVs, Annexin V^+^ MVs, and Annexin V^+^ RMVs compared to SLE-non-PAH patients. However, PAH patients with inactive/mild SLE only exhibited elevations in Annexin V^+^ MVs, RMVs, and Annexin V^+^ RMVs. In the SLE-PAH patients, EMVs were positively correlated with pulmonary arterial systolic pressure, while PMVs and EMVs were positively correlated with right ventricular diameter. Moreover, the receiver operating characteristic curve indicated that Annexin V^+^ MVs, LMVs, PMVs, RMVs, EMVs and Annexin V^+^ RMVs can predict the presence of PAH in SLE patients. Importantly, multivariate logistic regression analysis showed that circulating levels of LMVs or RMVs, anti-nRNP antibody, and serositis were independent risk factors for PAH in SLE patients. Finally, our findings reveal that specific subgroups of circulating MVs contribute to the hypercoagulation state and the severity of SLE-PAH. Higher plasma levels of LMVs or RMVs may serve as biomarkers for SLE-PAH.

## Introduction

Systemic lupus erythematosus (SLE) is a complex autoimmune disease affecting multiple organs with various clinical manifestations. Pulmonary arterial hypertension (PAH) is a serious complication of SLE, leading to death due to progressive right heart failure (1). PAH is characterized by increased pulmonary vascular resistance caused by different etiologies and pathogeneses (1, 2). Connective tissue diseases (CTDs) are the important cause of PAH. Systemic sclerosis (SSc)-associated PAH (SSc-PAH) is the most common type of CTD-PAH in Caucasians (2). However, SLE is the most common CTD-associated PAH in Chinese (2). Lack of specific clinical symptoms makes the early diagnosis of PAH difficult. PAH is an independent prognostic factor of SLE, as SLE-PAH patients experience significantly worse clinical outcomes than SLE-non-PAH patients (3). Without appropriate treatment, SLE-PAH patients face a tragically short average survival time of only two years from the onset of PAH (4). Thus, improving the prognosis of SLE-PAH patients remains as a critical challenge for rheumatologist. Although the pathogenesis of SLE-associated PAH still remains unclear, emerging evidence suggests the potential involvement of hypercoagulation and thrombosis in its development (5). However, the unclear risk-benefit profile of anticoagulantion therapy in SLE-PAH patients without antiphospholipid antibodies poses a challenge for treatment decisions (6–8). Therefore, better understanding of coagulation status in SLE-PAH patients may provide valuable insights into diseases prognosis and pave the way for novel therapeutic strategies.

Extracellular vesicles (EVs) are cellular membrane structures that are released from cells when they are stimulated or undergo different types of programmed cell death (9, 10). Among three types of EVs, microvesicles (MVs, 100–1000 nm in diameter) bud from the plasma membrane, exosomes (50–100 nm in diameter) are released from intracellular milieu, while apoptotic bodies are by-products of apoptotic cells (9–11). Studies from our and others have demonstrated that MVs may harbor proteins, lipids, nucleic acids, or multi-molecular complexes from the nucleus, cytoplasm, or plasma membrane of their parental cells (10, 12). Functionally, MVs can serve as novel mediators for intercellular communication within organs or across different organ systems (10). Among the diverse array of biological features MVs possess, their roles in procoagulation remain the most studied function, with implications for various human diseases. Our previous studies have shown that MVs from monocyte/macrophages, with cholesterol enrichment or stimulation by tobacco smoke extracts, have potent procoagulant activities due to their-associated phosphatidylserine (PS) and other procoagulant factors (13, 14). MV shedding process results in exposure of membrane PS, a phospholipid crucial for procoagulant activity by providing surface for assembly of prothrombin complex, thus promoting thrombus formation (13, 14). Annexin V, a calcium-dependent phospholipid-binding protein that can preferentially bind PS on EV surface, becomes a valuable tool for detecting MVs with procoagulant functions (13, 14). In addition, circulating MVs derived from platelets (PMVs), red blood cells (RMVs), leukocytes (LMVs), and endothelial cells (EMVs) (15) are also important in cardio-pulmonary diseases (16, 17).

Various studies from our and other teams have shown that MVs contribute to PAH, with elevated procoagulant conditions or even pulmonary embolism (18, 19). However, circulating MVs and their pathogenic involvement in PAH associated with SLE remain unexplored. Furthermore, the inconsistent survival advantage of anticoagulant therapy for SLE-PAH patients compared to iPAH and SSc-PAH (6–8) suggests that the underlying mechanisms of SLE-PAH may not be fully captured by previous studies. In the current study, we systematically analyzed the levels of circulating MVs from different cell types, such as LMVs, RMVs, PMVs, or EMVs, and procoagulant Annexin V^+^ MVs, using flow cytometry, to determine the potential involvement of circulating MVs in pathogenesis of PAH in SLE patients. Through this study, we sought to provide evidence for the involvement of circulating MVs in development of PAH in patients with SLE, paving the way for new therapeutic targets or diagnostic tools.

## Material and methods

### Study population and data collection

Subjects were enrolled from August 2019 to December 2021 at the Department of Rheumatology in Tianjin Medical University General Hospital. A total of 18 SLE-PAH patients; 36 SLE-non-PAH patients with matched age, sex, and SLE disease activity (SLEDAI-2K) (24); and 36 healthy volunteers with matched age and sex were recruited to the study. All 54 SLE patients met the 2019 European League Against Rheumatism/American College of Rheumatology classification criteria for systemic lupus erythematosus (25). Seven PAH patients were diagnosed by right heart catheterization (RHC) with criteria of a mean pulmonary arterial pressure (mPAP) ≥ 25 mmHg at rest, a pulmonary capillary wedge pressure (PCWP) ≤ 15 mmHg, and pulmonary vascular resistance (PVR) > 3 Wood Units (26); while the other 11 PAH patients were diagnosed by pulmonary arterial systolic pressure (PASP) > 50 mmHg with ultrasonic cardiogram (UCG). Other causes of PAH (27) were excluded to ensure that PAH was attributable to SLE. SLE patients often had interstitial lung disease (ILD). When forced vital capacity was less than 60% predicted or, fibrosis lesion was large than 1/3 lung fields on high-resolution computed tomography (HRCT), the patients were also excluded to avoid ILD-associated PAH (2). PASP, as estimated by tricuspid regurgitation in UCG of SLE-non-PAH patients, could not be more than 30 mmHg. This study was approved by the Institutional Medical Ethics Review Boards of Tianjin Medical University General Hospital and informed consent was obtained from all study participants.

The following clinical data were collected from medical records: demographics, medical history, symptoms and syndromes, RHC results, laboratory tests (routine examinations of blood, urine and stool, phase microscope for hematuria, liver and renal function, coagulation function, indexes correlated with immune status, inflammatory markers, 24 hours proteinuria), and imageological examination (UCG, abdominal ultrasound, pulmonary HRCT, and magnetic resonance imaging of brain).

### Sample preparation

All study subjects fasted for more than 8 h before venous blood samples were collected between 7 a.m. and 9 a.m. Three milliliters of peripheral blood was collected in a Vacutainer™ (BD Biosciences, San Jose, CA, USA) containing 3.2% sodium citrate (volume of anticoagulation: volume of blood = 1:9) and processed within 2 h. The sample was centrifuged at 120 x *g* for 20 min to obtain the platelet-rich plasma supernatant. This was then centrifuged at 1,500 x *g* for 20 min and the platelet-poor plasma supernatant was collected. The platelet-poor plasma was further centrifuged at 13,000 x *g* for 3 min to obtain cell-free plasma (CFP). All centrifugations were performed at 20 °C and no more than one freeze-thaw cycle was permitted.

### Flow Cytometry Analysis

A panel of cell-specific monoclonal antibodies was used to label MVs originating from leukocytes (CD45), erythrocytes (CD235a), platelets (CD41a), and endothelial cells (CD144). Among the fluorescent antibody reagents, CD45-Brilliant Violet 650 (BV650), CD41a-fluorescein isothiocyanate (FITC), CD235a-Brilliant Violet 421 (BV421), CD144-Alexa Fluor 647, and their isotype-matched control conjugates were procured from BD Biosciences (San Jose, CA, USA). Annexin V–phycoerythrin (PE) was obtained from BioLegend (San Diego, CA, USA), and the negative control abolished the interaction of Annexin V with PS by the addition of ethylene diamine tetraacetic acid. The number of MVs were determined using high-sensitivity flow cytometry (BD LSRFortessa™ Cell Analyzer, BD Biosciences) following a previously described standardization method. Briefly, the upper limit of the MVs quadrant gate was established at 1 μm using standard microbeads that measured 0.5, 0.9, and 3 µm in diameter. The lower limit was determined with a threshold at side-scatter 200 nm (Figure 1A, B). Fifty microliters of CFP was incubated with the appropriate amount of antibodies of the parental cell type for 30 min. The mixtures were then incubated with 5 μL Annexin V and 50 μL 2×Annexin V binding buffer (BD Biosciences) for 15 min. All incubations were performed at 20 °C in the dark. After incubation, 10 μL count beads (Spherotech, Lake Forest, IL, USA) were added to the sample to calculate the absolute concentration of MVs, and all the samples were diluted to 500 μL with phosphate buffered saline (PBS) before analysis with flow cytometry. The absolute MV counts were determined using the following equation: (A/B) ×(C/D) = number of MVs per microliter. In this computational formula, A was the number of events for the test sample, B was the number of events for the count fluorescent beads, C was the total number of count fluorescent beads per 10 μL, and D was the volume of test sample initially used. All the buffers used in the MVs detection were filtered with 0.1 μm filters (Sartorius, Göttingen, Germany) to avoid contamination from non-cellular particles. Prior to use, the inner and outer tubes of the cytometer were completely cleaned until the background detection was fewer than 80 events/s. Sample analysis was run at a low speed and collection stopped at 60s.

**Figure 1.**
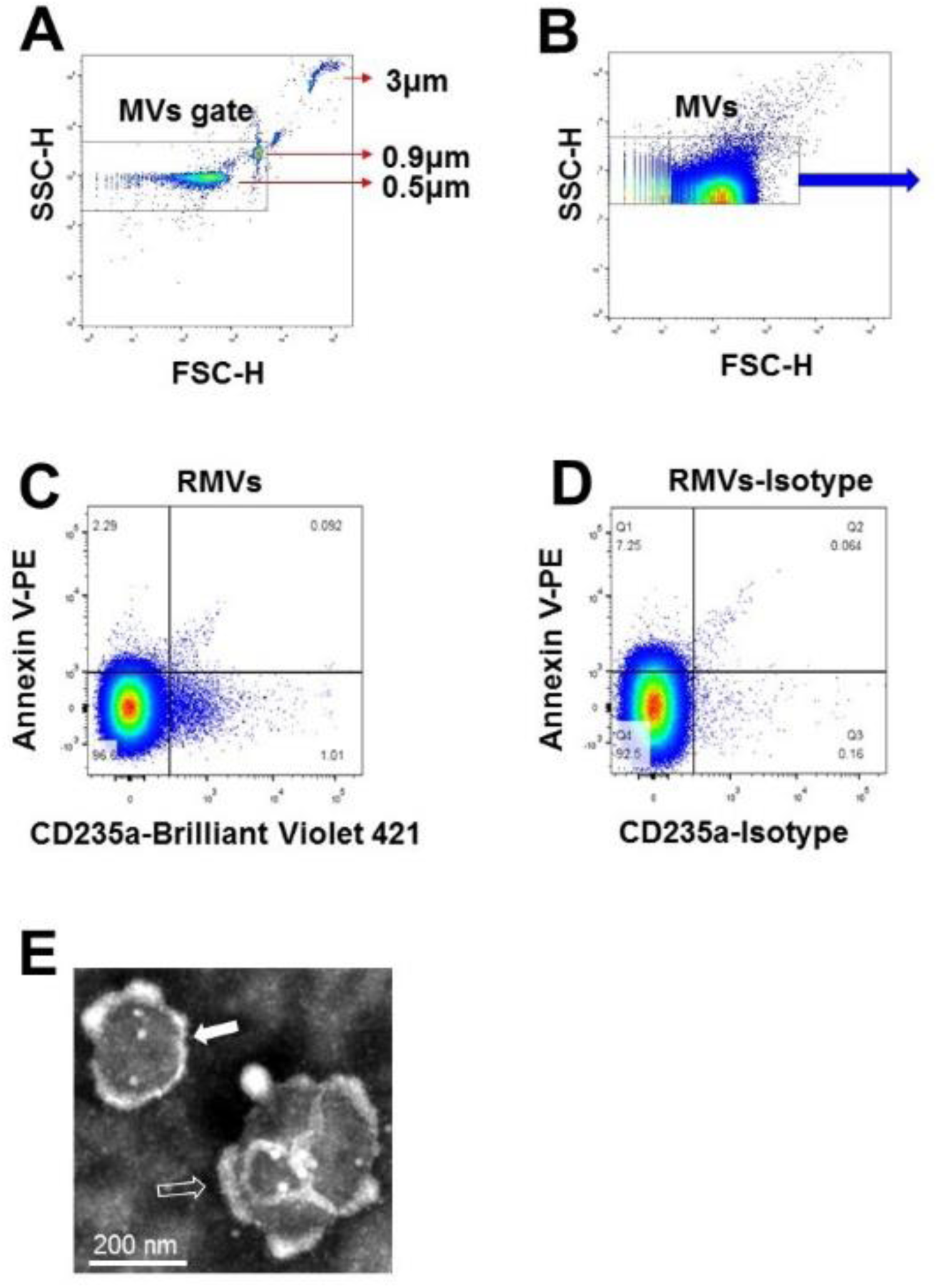
Characterization of MVs in plasma of SLE-PAH patients. **(A)**. Microbeads of 0.5, 0.9, and 3µm in diameter were used for gating MVs in flow cytometry analysis. **(B)**. MVs were gates for further analysis. **(C)** & **(D)**. Flow cytometry of MVs and their isotypes: erythrocytes-derived MVs. **(E)**. Transmission electron microscopy of isolated MVs from plasma of SLE patients. (Filled arrow demonstrated single MVs, black arrow indicated the clumped MVs).

### Transmission Electron Microscopy

Six milliliters of fresh CFP was centrifuged at 100,000 x *g* for 1 h at 4 °C to obtain the MVs sediment, which was then suspended in 60 μl PBS buffer to acquire pure MVs. A 150 Mesh carbon membrane/formvar membrane-coated grid was placed on a 30 μl MVs sample drop for 2 min at 20 °C, and a filter paper was used to remove excess liquid. The sample was negatively stained by placing the grid on a 30 μl drop of phosphotungstic acid (pH 6.5) for 2 min followed by blotting and air drying. Digital images were obtained using a Hitachi HT-7700 transmission electron microscope (Tokyo, Japan). As shown in Figure 1D, the MVs were separate from each other and had a spherical-shaped double membrane structure.

### Statistical analysis

The data with normal distribution were described as mean ± standard deviation (SD), and the median values were used to describe data with no normal distribution. The independent sample t-test was used to compare data for two groups, and the Mann-Whitney U-test was used if the data were not normally distributed. One-way analysis of variance, or the Kruskal-Wallis test for measurement data of abnormal distribution or unequal variances, was used to determine whether the variables in the three groups had significant differences. Receiver operating characteristic (ROC) curves were used to detect the capacity of circulating MVs subgroups to discriminate SLE-PAH patients from SLE-non-PAH patients. Conditional logistic regression analysis was used to explore the risk factors for the PAH complication in SLE patients. Spearman’s rank correlation coefficient was used to examine the strength of the relationship between the MVs subgroups and the variables measured by UCG. All tests were two-sided, and *p* < 0.05 was considered statistically significant. Statistical analyses were performed using SPSS 25.0 for Windows (SPSS Inc., IBM Corp., Armonk, NY, USA)

## Results

### Demographics and baseline characteristics of all participants

In this study, total of 18 SLE-PAH patients (one male) were enrolled, including 4 patients with inactive SLE (SLEDAI-2K<4), 4 patients with mild SLE (5≤SLEDAI-2K<10), 3 patients with moderate SLE (10≤ SLEDAI-2K<15), and 7 patients with severe SLE (SLEDAI-2K>15). In addition, 36 SLE non-PAH patients matched with age, sex, and SLE activity, and 36 healthy controls (HCs) matched with age and sex, were included. Hematology analyses showed that red blood cells (RBC), hemoglobin (HGB), platelet (PLT), and hematocrit (HCT) levels were decreased in SLE patients with or without PAH as compared with HCs, while the indexes of RBC distribution width (RDW), coefficient of variation of RDW (RDW-CV), and standard deviation of RDW (RDV-SD) were increased (Table 1). Compared with SLE-non-PAH patients, RDW-CV and RDV-SD, uric acid level, and anti-nRNP antibody positive rate were higher in SLE-PAH patients. Analysis of coagulation function showed increased fibrinogen levels, prothrombin time (PT), and PT-international normalized ratio (PT-INR) in SLE-PAH patients who received a higher rate of anticoagulant treatment. Among the clinical manifestations of SLE, patients with SLE-PAH were more likely to have serositis and Raynaud’s phenomenon. UCG indicated that SLE-PAH patients had higher pulmonary artery systolic pressure (PASP), right ventricular diameter (RVD), and proportion of pericardial effusion, which were consistent with the patients’ condition. The other parameters were comparable between the SLE with and without PAH groups. The detail information about clinical and laboratory characteristics are shown in Table 2.

**Table 1.** Demographics and hematology analysis in SLE patients and healthy controls.

| Variables | Healthy controls<br>(n=36) | SLE-non-PAH<br>(n=36) | SLE-PAH<br>(n=18) | p-value | p-value for two groups |  |  |
| --- | --- | --- | --- | --- | --- | --- | --- |
|  |  |  |  |  | HC vs.<br>SLE-non-PAH | HC vs.<br>SLE-PAH | SLE-non-PAH vs.<br>SLE-PAH |
| Demographics |  |  |  |  |  |  |  |
| Age (years) | 46.3±13.2 | 46.6±15.9 | 47.4±15.8 | 0.949 | - | - | - |
| Gender (F/M) | 34/2 | 34/2 | 17/1 | 1.000 | - | - | - |
| Hematology analysis |  |  |  |  |  |  |  |
| WBC, ×10<sup>9</sup>/L | 5.6 (5.1, 6.9) | 5.2 (3.7, 6.7) | 6.4 (4.1, 11.2) | 0.116 | - | - | - |
| RBC, ×10<sup>12</sup>/L | 5.0 (4.7, 5.4) | 3.6 (3.1, 4.0) | 3.7 (3.2, 4.3) | <0.001 | <0.001 | <0.001 | 0.732 |
| HGB, g/L | 137 (133, 141) | 111 (90, 117) | 99 (93, 128) | <0.001 | <0.001 | <0.001 | 0.811 |
| PLT, ×10<sup>9</sup>/L | 256 (211, 296) | 202 (136, 259) | 188 (100, 294) | 0.001 | 0.002 | 0.012 | 0.666 |
| HCT, % | 42.7 (41.5, 43.6) | 32.4 (28.1, 36.2) | 32.8 (29.4, 39.8) | <0.001 | <0.001 | <0.001 | 0.388 |
| RDW-CV, % | 12.8 (12.2, 13.6) | 13.8 (12.8, 16.0) | 15.8 (14.3, 18.5) | <0.001 | 0.002 | <0.001 | 0.012 |
| RDW-SD, fL | 40.9 (39.7, 42.9) | 44.1 (41.3, 50.7) | 53.3 (47.4, 57.7) | <0.001 | 0.002 | <0.001 | <0.001 |
| PDW, fL | 12.8 (1170, 13.8) | 11.8 (10.6, 13.1) | 10.7 (9.0, 16.0) | 0.096 | - | - | - |
| P-LCR, % | 27.8 (26.0, 29.1) | 28.3 (25.0, 34.9) | 23.6 (17.7, 36.4) | 0.412 | - | - | - |

Data are median (interquartile range) unless otherwise stated. WBC: white blood cell; RBC: red blood cell; HGB: hemoglobin; PLT: platelet; HCT: hematocrit; RDW-CV: coefficient of variation of red blood cell distribution width; RDW-SD: standard deviation of red blood cell distribution width; PDW: platelet distribution width; P-LCR: platelet-large cell ratio. Bold font indicates statistical significance.

**Table 2.** Clinical characteristics in SLE patients with and without PAH.

| Variables | SLE-non-PAH<br>(n=36) | SLE-PAH<br>(n=18) | p-value |
| --- | --- | --- | --- |
| SLE duration, months (median, quartile) | 48.0 (2.6, 117.0) | 79.0 (11.8, 240.0) | 0.053 |
| SLEDAI | 12±7 | 10±8 | 0.381 |
| Laboratory parameters |  |  |  |
| CRP, mg/dl | 0.26 (0.18, 1.35) | 0.67 (0.40, 1.20) | 0.066 |
| C3, mg/dl | 53.4±19.5 | 65.8±27.5 | 0.063 |
| C4, mg/dl | 13.5±6.0 | 15.2±6.4 | 0.051 |
| Anti-dsDNA positive (n, %) | 19 (52.8%) | 7 (38.9%) | 0.336 |
| Anti-nRNP positive (n, %) | 14 (38.9%) | 13 (72.2%) | **0.021** |
| ALB, g/L | 31.9±6.0 | 31.4±4.4 | 0.772 |
| ALT, U/L | 20 (13, 42) | 17 (13,35) | 0.890 |
| Creatinine, µmol/L | 54 (41, 61) | 64 (46, 99) | 0.189 |
| Uric acid, µmol/L | 309.8±113.9 | 413.8±160.9 | **0.009** |
| PT, seconds | 11.3 (10.7, 11.7) | 11.8 (11.3, 25.1) | **0.007** |
| PT-INR (median, quartile) | 1.03 (0.98, 1.07) | 1.08 (1.03, 2.27) | **0.007** |
| APTT, seconds (median, quartile) | 29.5 (27.3, 34.8) | 33.0 (29.2, 37.6) | 0.104 |
| TT, seconds (median, quartile) | 21.0 (19.4, 24.2) | 20.2 (19.4, 22.1) | 0.304 |
| Fibrinogen, g/L (median, quartile) | 2.56 (2.31, 2.95) | 3.41 (2.45, 4.02) | **0.040** |
| D-Dimer, ng/mL (median, quartile) | 742 (379, 2404) | 735(482, 1208) | 0.474 |
| SLE manifestations (n, %) |  |  |  |
| Nervous involvement | 3 (8.3%) | 1 (5.6%) | 0.713 |
| Renal involvement | 21 (58.3%) | 9 (50.0%) | 0.561 |
| Haematological involvement | 29 (80.6%) | 13 (72.2%) | 0.487 |
| Rash | 11 (30.6%) | 3 (16.7%) | 0.177 |
| Arthritis | 5 (13.9%) | 2 (11.1%) | 0.775 |
| Serositis | 11 (30.6%) | 12 (66.7%) | **0.011** |
| Raynaud phenomenon | 1 (2.8%) | 4 (22.2%) | **0.003** |
| Gastrointestinal involvement | 7 (19.4%) | 1 (5.6%) | 0.176 |
| UCG |  |  |  |
| PASP, mmHg | 22 (18, 26) | 61 (50, 73) | **<0.001** |
| RV diameter, mm | 35 (32, 37) | 41 (37,49) | **0.008** |
| Pericardial effusion (n, %) | 3 (8.6%) | 7 (38.9%) | **0.008** |
| 6MWD*, m (median, quartile) | - | 365 (0, 414) | - |
| BNP, ng/L (median, quartile) | 26.7 (14.8, 127.3) | 390.0 (82.7, 912.5) | **0.003** |
| Medications |  |  |  |
| Glucocorticoids (n, %) | 22 (61.1%) | 13 (72.2%) | 0.420 |
| Immunosuppressants (n, %) | 13 (36.1%) | 8 (44.4%) | 0.554 |
| Anticoagulant | 1 (3.3%) | 9 (50%) | **<0.001** |

CRP: C-reactive protein; C3/4: Complement 3/4; anti-dsDNA: anti-double strand DNA antibody; anti-RNP: anti-nuclear ribonucleoprotein antibody; ALB: Albumin; ALT: Alanine transaminase; PT: prothrombin time; PT-INR: prothrombin time- International Normalized Ratio; APTT: activated partial thromboplastin time; TT: thrombin time; PASP: pulmonary artery systolic pressure; RV: right ventricle; \*6MWD: 6-min walk distance, 0m was assigned to patients who were admitted to the hospital with symptoms of obvious right heart failure and were unable to tolerate the test; BNP: brain natriuretic peptide. Bold font indicates statistical significance.

### Circulating MVs in SLE patients with and without PAH

When we compared all circulating MVs among SLE patients with or without PAH, and healthy controls, we found significantly increased plasma levels of LMVs (CD45^+^LMVs), PMVs (CD41a^+^PMVs), RMVs (CD235a^+^RMVs), EMVs (CD144^+^EMVs), and Annexin V^+^MVs in SLE patients with or without PAH as compare to healthy controls (Figure 2A-E). In addition, the dual stainings of Annexin V^+^ LMVs, Annexin V^+^ PMVs, Annexin V^+^ RMVs, and Annexin V^+^ EMVs were also elevated in both SLE patient groups as comparted to those in HC subjects (Figure 2F-I), indicating the increased generation of MVs from leukocytes, platelets, RBCs, and vascular endothelium in SLE patients no matter if they have PAH or not.

**Figure 2.**
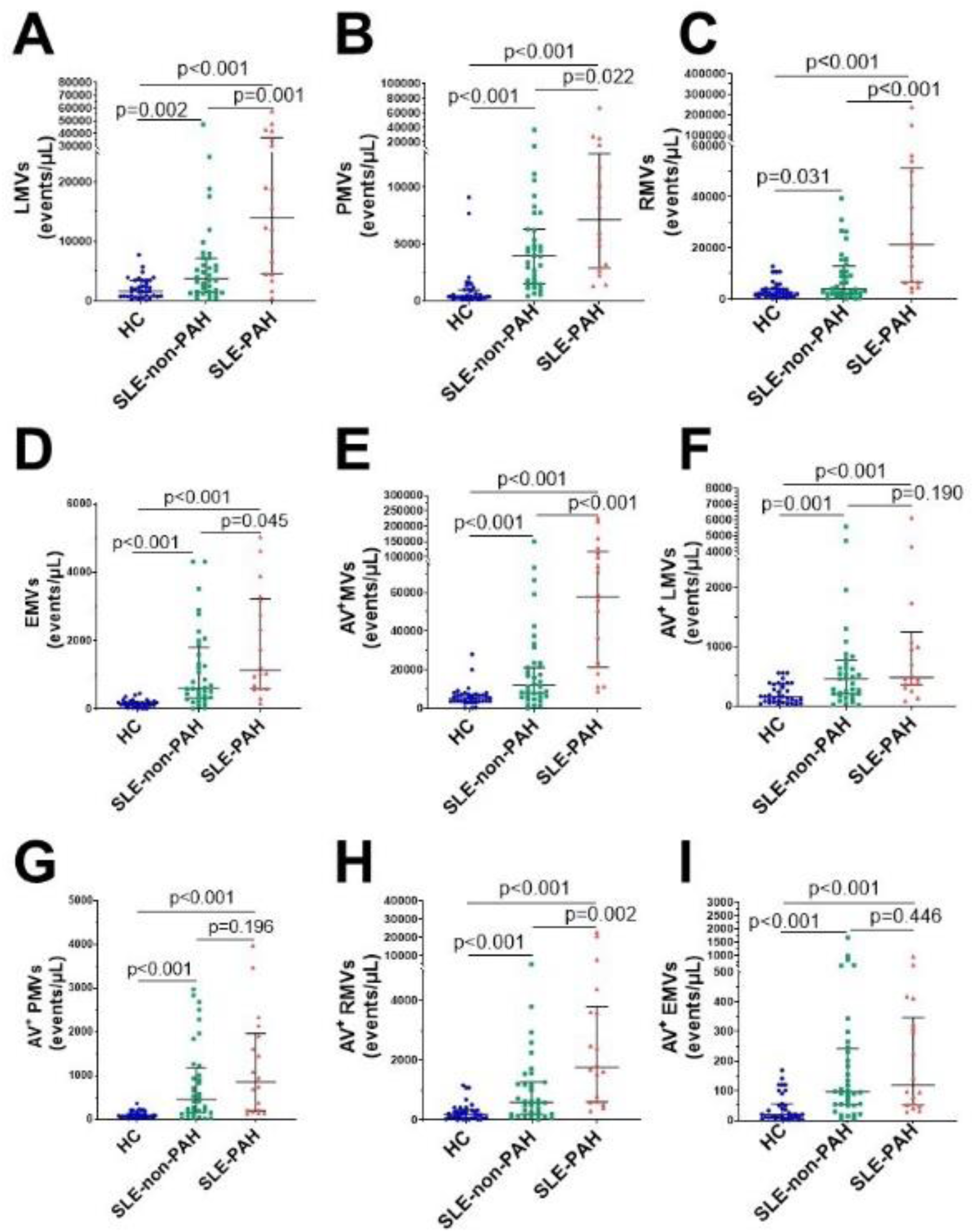
Circulating MVs in SLE patients with or without PAH, compared with healthy control groups. **(A)** Statistical results of single-labeled LMVs; **(B)** PMVs; **(C)** RMVs; **(D)** EMVs; **(E)** Annexin V^+^MVs in SLE patients with/without PAH, and HC group. **(F)** Statistical results of dual-labeled Annexin V^+^LMVs; **(G)** Annexin V^+^PMVs; **(H)** Annexin V^+^RMVs; **(I)** Annexin V^+^EMVs in SLE patients with/without PAH, and HC group.

To further explore the relationship MVs and PAH in SLE patients with or without PAH, we compared single-labeled MVs, i.e. LMVs, PMVs, RMVs, EMVs, and MVs, as well as the dual-labeled MVs, i.e. Annexin V^+^ LMVs, Annexin V^+^ PMVs, Annexin V^+^ RMVs, and Annexin V^+^ EMVs, between the SLE-PAH and SLE-non-PAH groups (Table 3). All single-labeled MVs (LMVs, PMVs, RMVs, EMVs and Annexin V^+^ MVs), and the dual-labeled MVs (Annexin V^+^ RMVs) were significantly increased in SLE-PAH group as comparted to those in SLE-non-PAH group (Figure 2), suggesting the potential role of circulating MVs in PAH. In order to eliminate the impact of SLE disease severity, patients were further divided into an inactive/mild SLE group (SLEDAI-2K<10) and a moderate/high SLE group (SLEDAI-2K≥10). Interestingly, the plasma levels of Annexin V^+^MVs, RMVs, and Annexin V^+^ RMVs were elevated in SLE-PAH patients with inactive/mild SLE activities; while the SLE-PAH patients with moderate/high SLE activities have increased plasma levels of LMVs, PMVs, RMVs, Annexin V^+^MVs, and Annexin V^+^ RMVs (Table 4, Figure 3).

**Figure 3.**
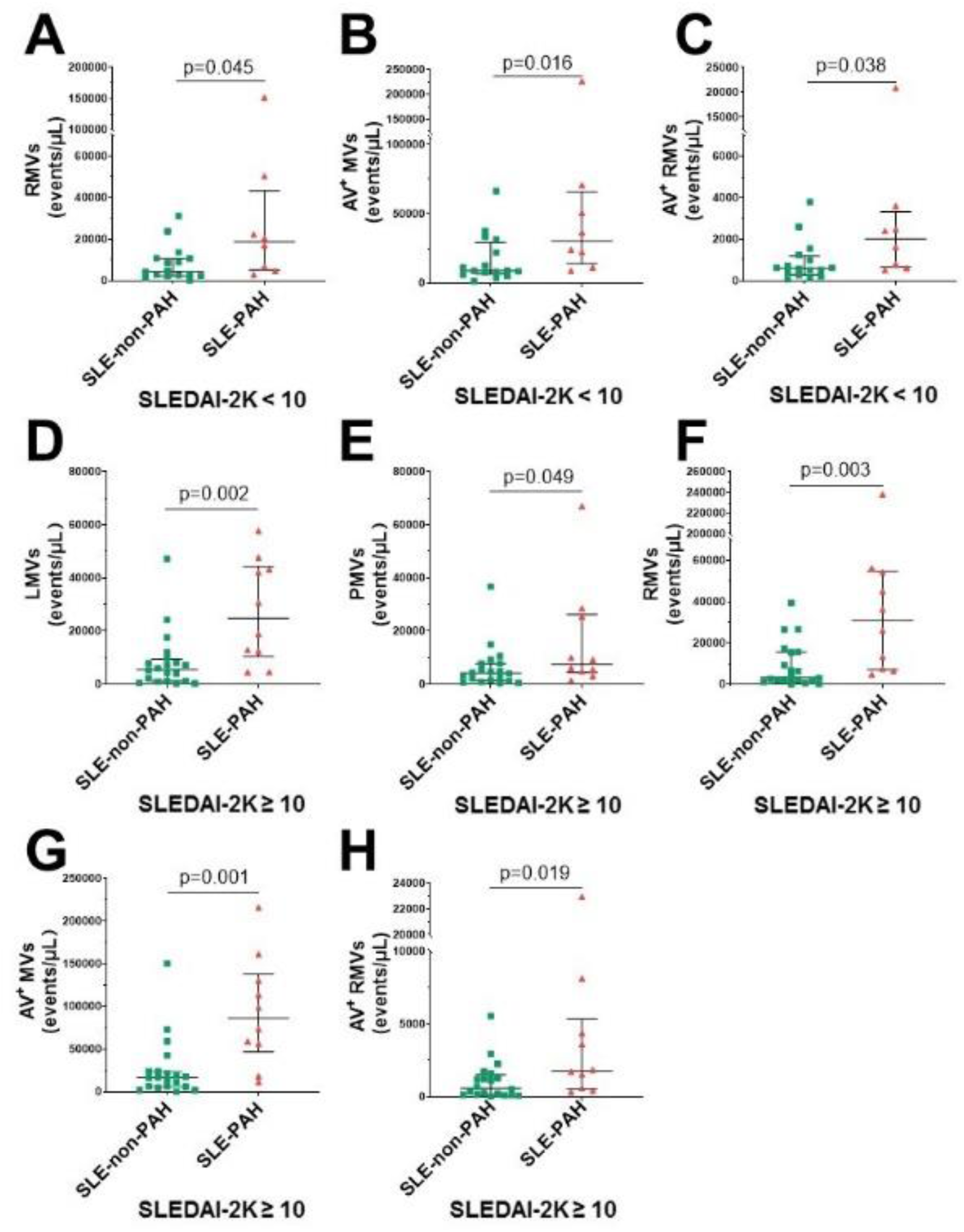
Circulating MVs in different SLE activity. **(A)** SLEDAI-2K<10 SLE-PAH patients (n=8) compared with SLE-non-PAH patients (n=16) statistical results of RMVs; **(B)** Annexin V^+^MVs; **(C)** Annexin V^+^RMVs. **(D)** SLEDAI-2K≥10 SLE-PAH patients (n=10) compared with SLE-non-PAH patients (n=20) statistical results of LMVs; **(E)** PMVs. **(F)** RMVs; **(G)** Annexin V^+^MVs; **(H)** Annexin V^+^RMVs.

**Table 3.** Circulating MVs and their cellular origin in different groups of subjects.

| Origin | Healthy controls<br>(n=36) | SLE-non-PAH<br>(n=36) | SLE-PAH<br>(n=18) | p-value |  |  |
| --- | --- | --- | --- | --- | --- | --- |
|  |  |  |  | SLE-non-PAH<br>vs. HC | SLE-PAH<br>vs. HC | SLE-non-PAH<br>vs. SLE-PAH |
| Total procoagulant |  |  |  |  |  |  |
| Annexin V<sup>+</sup> MVs | 5263 (3778, 7481) | 11959 (6029, 23610) | 57783 (21319, 117546) | <0.001 | <0.001 | <0.001 |
| Leukocytes |  |  |  |  |  |  |
| LMVs | 1595 (793, 3457) | 3697 (1450, 7216) | 14285 (4512, 36833) | 0.002 | <0.001 | 0.001 |
| Annexin V<sup>+</sup> LMVs | 158 (68, 370) | 459 (198, 777) | 483 (355, 1250) | 0.001 | <0.001 | 0.190 |
| Platelets |  |  |  |  |  |  |
| PMVs | 401 (297, 997) | 3965 (1531, 6296) | 7129 (2920, 12965) | <0.001 | <0.001 | 0.022 |
| Annexin V<sup>+</sup> PMVs | 84 (55,134) | 453 (152, 1169) | 848 (197, 1963) | <0.001 | <0.001 | 0.196 |
| Red blood cells |  |  |  |  |  |  |
| RMVs | 2395 (1187, 4164) | 3909 (2027, 12900) | 21365 (6643, 51282) | 0.031 | <0.001 | <0.001 |
| Annexin V<sup>+</sup> RMVs | 187 (70, 330) | 598 (196, 1262) | 1769 (608, 3804) | <0.001 | <0.001 | 0.002 |
| Endothelial cells |  |  |  |  |  |  |
| EMVs | 163 (109, 203) | 615 (327, 1808) | 1143 (604, 3219) | <0.001 | <0.001 | 0.045 |
| Annexin V<sup>+</sup> EMVs | 20 (10, 55) | 97 (52, 242) | 120 (52, 346) | <0.001 | <0.001 | 0.446 |

Bold font indicates statistical significance.

**Table 4.** Circulating MVs in SLE patients with different severity.

| Origin | SLEDAI<10 |  | p-value | SLEDAI≥10 |  | p-value |
| --- | --- | --- | --- | --- | --- | --- |
|  | SLE-non-PAH<br>(n=16) | SLE-PAH<br>(n=8) |  | SLE-non-PAH<br>(n=20) | SLE-PAH<br>(n=10) |  |
| Total procoagulant |  |  |  |  |  |  |
| Annexin V<sup>+</sup> | 8887 (6420, 29197) | 30323 (14045, 65717) | **0.016** | 16493 (5925, 23610) | 86113 (46839, 137743) | **0.001** |
| Leukocytes |  |  |  |  |  |  |
| LMVs | 3067 (1580, 5051) | 9371 (2084, 18232) | 0.093 | 5574 (1204, 9416) | 24803 (10415, 44269) | **0.002** |
| Annexin V<sup>+</sup> LMVs | 421 (229, 780) | 477 (193, 1481) | 0.610 | 489 (142, 777) | 711 (355, 1672) | 0.183 |
| Platelets |  |  |  |  |  |  |
| PMVs | 3664 (1605, 5892) | 5755 (2347, 11406) | 0.264 | 4161 (1249, 7746) | 7597 (4429, 26261) | **0.049** |
| Annexin V<sup>+</sup> PMVs | 412 (108, 1152) | 622 (155, 2021) | 0.383 | 453 (202, 1199) | 1098 (208, 1963) | 0.286 |
| Red blood cells |  |  |  |  |  |  |
| RMVs | 4514 (2382, 10724) | 18694 (5168, 43325) | **0.045** | 3336 (1510, 15559) | 31062 (7201, 54661) | **0.003** |
| Annexin V<sup>+</sup> RMVs | 598 (291, 1178) | 2008 (660, 3319) | **0.038** | 590 (119, 1497) | 1769 (541, 5322) | **0.019** |
| Endothelial cells |  |  |  |  |  |  |
| EMVs | 536 (327, 825) | 1003 (390, 2734) | 0.214 | 1174 (270, 2221) | 2049 (669, 3368) | 0.155 |
| Annexin V<sup>+</sup> EMVs | 71 (35, 163) | 96 (63, 268) | 0.192 | 126 (64, 549) | 261 (45, 413) | 0.948 |

Bold font indicates statistical significance.

### Correlation between circulating MVs and severity of PAH

To explore the relationship between MVs and PAH, we analyzed the correlation analysis between each MVs subgroup and UCG parameters, as only 7 SLE-PAH patients had RHC results at the time of sample collection. The results showed that LMVs, RMVs, EMVs, and Annexin V^+^MVs, were positively correlated with PASP in the combined groups of all SLE patients (n=54). Further analysis of SLE-PAH patients showed positive correlation between EMVs and PASP (r=0.274, p=0.045). In addition, plasma levels of PMVs and EMVs were positively correlated with RVD. These results demonstrated significant relationship between platelets and endothelial cell-derived MVs and PAH severity (Table 5).

**Table 5.**
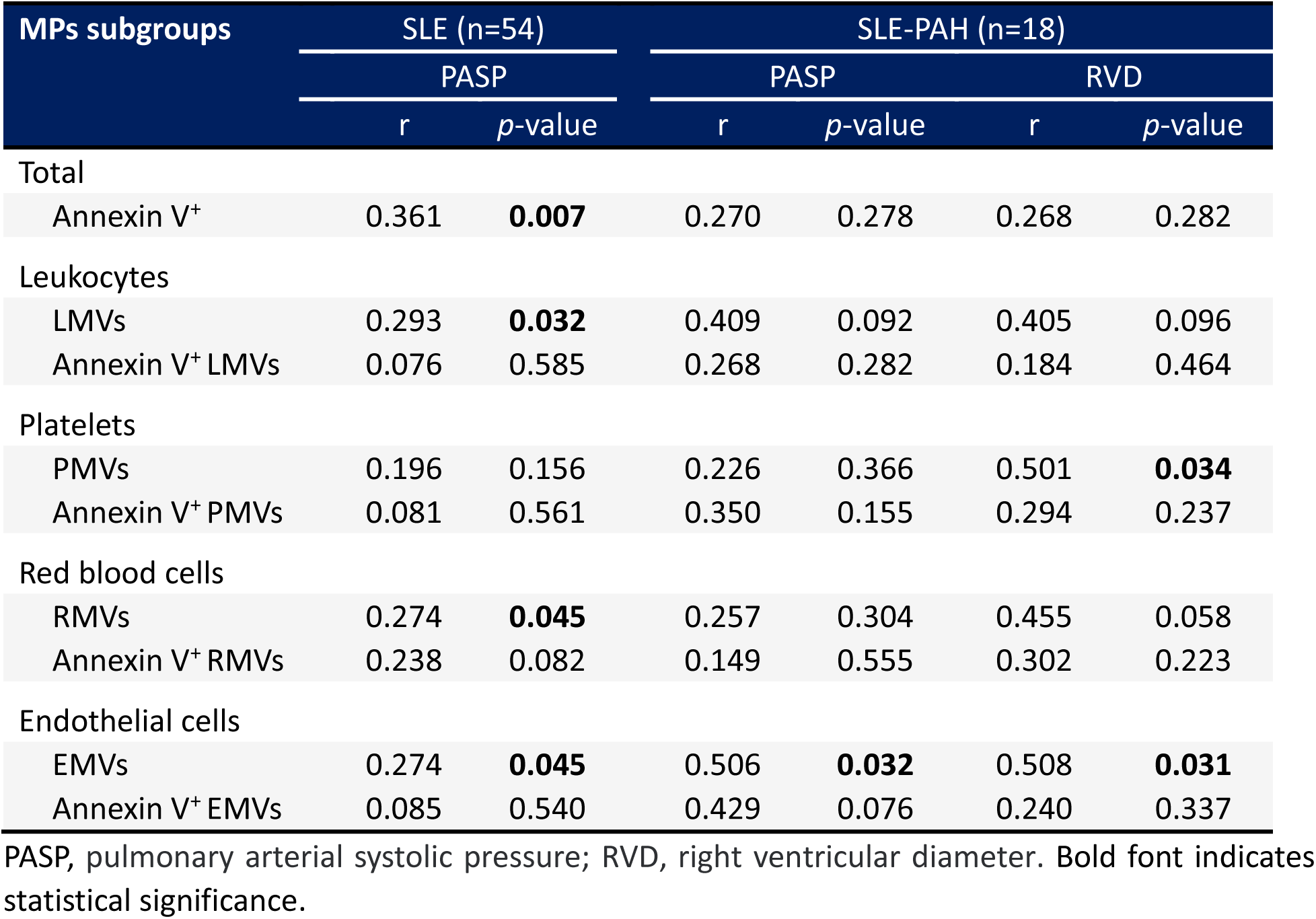
Correlation between MVs subgroups and UCG parameters in all SLE or SLE-PAH patients.

### Receiver operating characteristic analysis of Circulating MVs in SLE patients with PAH

To examine the specificity and sensitivity of circulating MVs, the ROC analyses found that plasma levels of LMVs, PMVs, RMVs, EMVs, Annexin V^+^MVs, and the dual-labeled Annexin V^+^ RMVs could clearly distinguish SLE-PAH from SLE-non-PAH (Figure 4A-F). Among the characteristics of circulating MVs, Annexin V^+^ MVs was the best predictor of PAH in patients with SLE for both specificity and sensitivity (Figure 4E, Table 6), suggesting the importance of Annexin V-positive, PS-exposing MVs in the development of PAH in SLE patients. In addition, LMVs are significant predictors of SLE-PAH (Table 6), suggesting that leukocyte-derived LMVs and their associated cago molecules play an important role in SLE-PAH development. Importantly, both RMVs and Annexin V^+^ RMVs predict SLE-PAH (Table 6), implicating RMV-associated PS in SLE-PAH pathogenesis. On the other hand, EMVs also significantly predict SLE-PAH (Table 6), indicating the involvement of endothelial damage in SLE-PAH pathogenesis.

**Figure 4.**
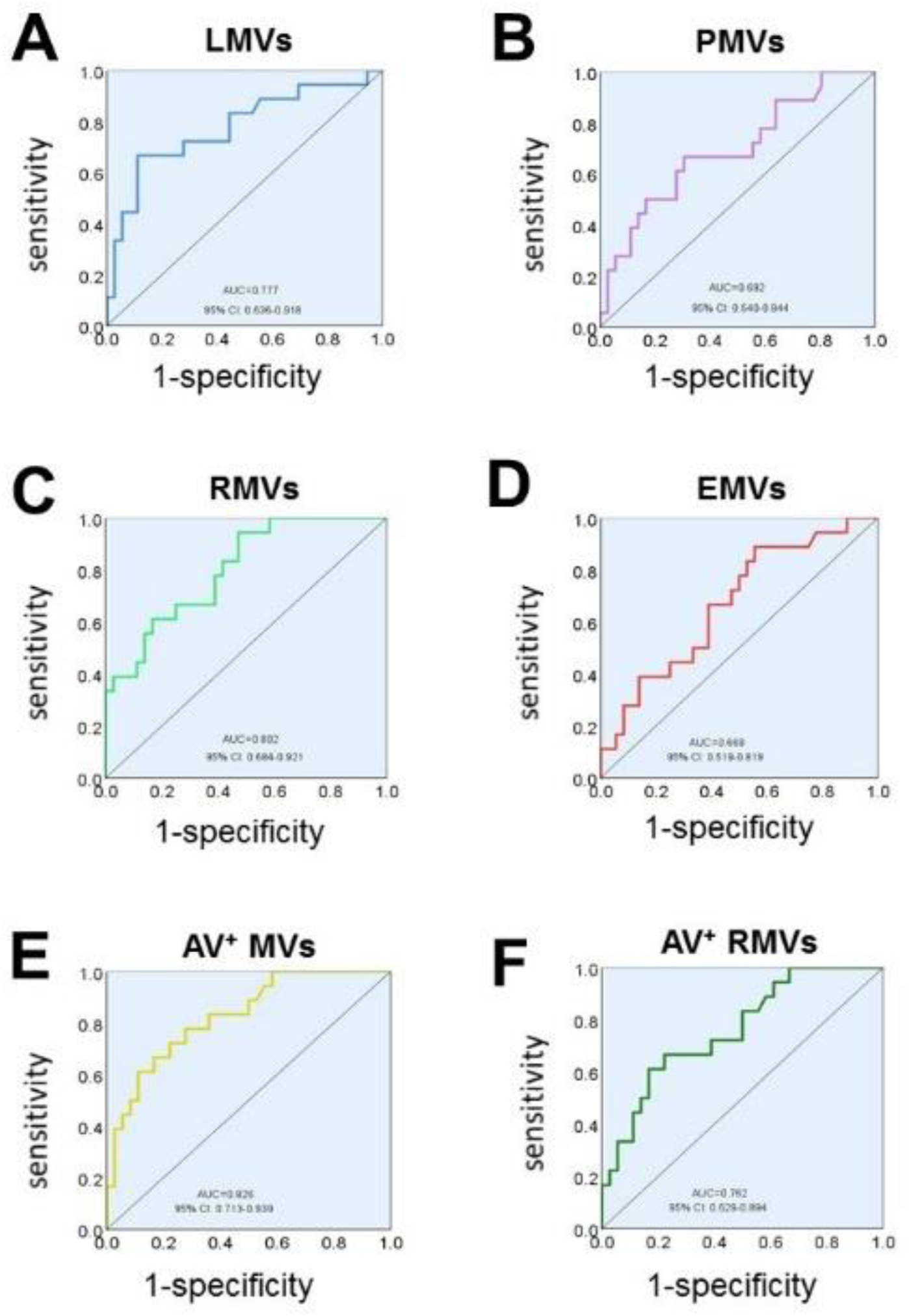
Receiver operating characteristic (ROC) analysis. ROC analysis shows the ability of LMVs **(A)**, PMVs **(B)**, RMVs **(C)**, EMVs **(D)**, Annexin V+MVs **(E)**, Annexin V+ RMVs **(F)** concentrations in discriminating SLE-PAH (n=18) from SLE-non-PAH (n=36).

**Table 6.**
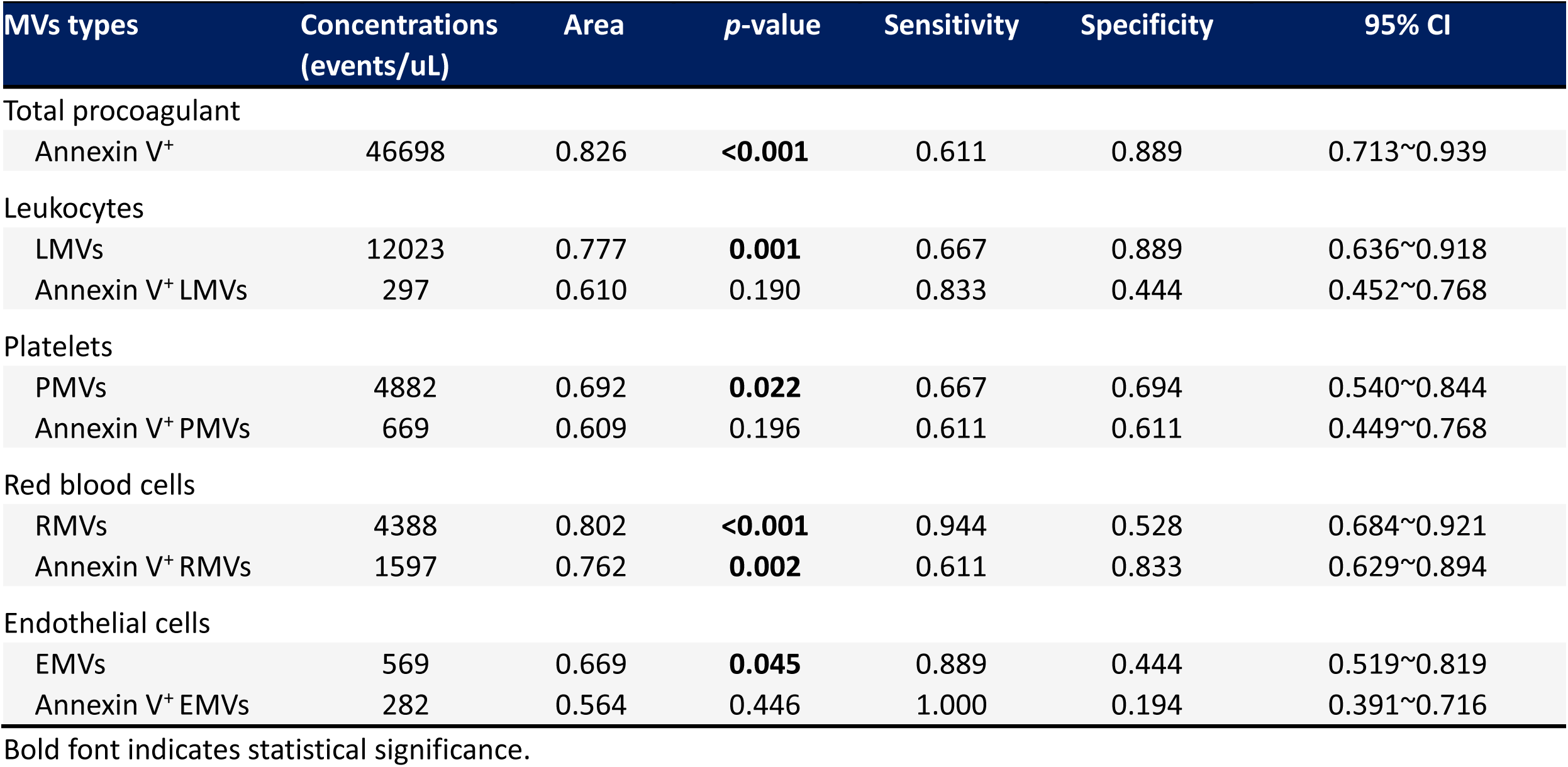
Circulating MVs concentrations for distinguish SLE-PAH from SLE-non-PAH.

### Associations between circulating MVs and PAH in SLE

To determine the prediction ability of circulating MVs and other associated clinical and laboratory indicators (based on variables in Table 1 and 2 and clinical implication) on PAH in SLE patients, we conducted univariate and multivariate logistic regression analyses (Table 7). Age, sex, and SLE disease activity were not included in the conditional logistic regression model as the groups were matched for them, and UCG parameters were not included because of their role in the inclusion and exclusion criteria for PAH. Anticoagulant was excluded because it was the treatment strategy based on a comprehensive assessment of the patient’s condition, and it was unrelated to the differential diagnoses. In univariate logistic regression analyses, the levels of LMVs (>12023 events/μL), PMVs (>4882 events/μL), RMVs (>4388 events/μL), EMVs (>569 events/μL), Annexin V^+^MVs (>46698 events/μL), and Annexin V^+^RMVs (>1597 events/μL), as well as RBC-SD, RBC-CV, UA, positive anti-nRNP antibody and serositis were risk factors of PAH. Most importantly, LMVs and RMVs, serositis, and positive anti-nRNP antibody were independently associated with the development of PAH in SLE patients (Table 7), indicating that LMVs and RMVs play an important role in development of PAH in SLE patients.

**Table 7.** Univariate and multivariate Logistic regression analysis for identifying PAH in SLE patients (n=54).

| Variables | Univariate analysis |  | Multivariate analysis |  |
| --- | --- | --- | --- | --- |
|  | Odds ratio (95% CI) | *p*-value | Odds ratio (95% CI) | *p*-value |
| LMVs >12023 events/μL | 16.000 (3.834-66.763) | **<0.001** | 7.376 (1.067-50.988) | **0.043** |
| PMVs >4882 events/μL | 4.545 (1.356-15.238) | **0.014** | - | |
| RMVs >4388 events/μL | 19.000 (2.280-158.336) | **0.006** | 31.297 (2.157-454.004) | **0.012** |
| EMVs >569 events/μL | 6.400 (1.279-32.027) | **0.024** | - | |
| Annexin V<sup>+</sup> MVs >46698 events/μL | 12.571 (3.080-51.315) | **<0.001** | - | |
| Annexin V<sup>+</sup> RMVs >1597 events/μL | 7.857 (2.161-28.568) | **0.002** | - | |
| RBC-SD | 1.122 (1.033-1.218) | **0.006** | - |  |
| RBC-CV | 1.267 (1.018-1.576) | **0.034** | - |  |
| Uric acid | 1.006 (1.001-1.012) | **0.019** | - |  |
| anti-nRNP positive | 4.086 (1.194-13.978) | **0.025** | 10.278 (1.365-77.381) | **0.024** |
| serositis | 4.545 (1.356-15.238) | **0.014** | 10.186 (1.375-75.473) | **0.023** |

RDW-CV: coefficient of variation of red blood cell distribution width; RDW-SD: standard deviation of red blood cell distribution width; anti-RNP: anti-nuclear ribonucleoprotein antibody. Bold font indicates statistical significance.

## Discussion

Circulating MVs have been reported to be associated with disease severity in SLE patients (28). In PAH, studies have investigated the differences of specific subsets of MVs in patients with iPAH, hPAH, and CTD-PAH (29, 30). However, no studies have investigated the circulating MVs in SLE-PAH patients. Here, we showed elevated circulating MVs, including LMVs, PMVs, RMVs, EMVs, Annexin V^+^MVs, and double-labeled Annexin V^+^RMVs, in SLE-PAH patients compared to SLE-non-PAH patients. The results suggest that these circulating MV subgroups are related to the pathologic conditions of PAH in patients with SLE.

As discussed above, studies from our the other groups have shown that Annexin V^+^MVs have procoagulant phospholipid activities due to their exposure of PS (31). In this study, we demonstrated that total Annexin V^+^MVs and Annexin V^+^ MVs from different circulating cells or endothelial cells were significantly increased in both SLE-non-PAH and SLE-PAH patients compared to HCs, indicating the involvement of MVs, particularly the MVs with PS exposure, in the development of SLE. Importantly, the levels of Annexin V^+^MVs in SLE-PAH patients were significantly higher than in SLE-non-PAH patients, suggesting the importance of procoagulant MVs in the development of PAH in SLE patients. Given the established role of Annexin V-positive MVs with PS exposure in procoagulation (31), these results support the pathologic involvement of MVs and the key role of a pro-thrombotic condition in the development of PAH in SLE.

RBCs are the most abundant circulating blood cells (32), and the primary contributors of circulating MVs (33). In our study, we demonstrated that RMVs emerged as the highest PS-exposing MVs among all cell types examined, highlighting the contribution of RMVs in the prothrombotic conditions in PAH development in SLE patients. It’s known that RBCs can release RMVs when erythrocytes encounter specific stimulations, such as shear stress, complement attack, pro-apoptotic stimulation, and injury (34). Previous studies demonstrate that changes in shear stress in PAH patients (35) are accompanied by abnormal morphology of erythrocyte (36). Abnormal morphology of RBC can be found in RBC with increased RDW, while in the most representative disease of increased RDW, sickle cell disease, both RMV release and PS exposure in erythrocytes are increased, accompanied by increased coagulation activation marker (37). The number of RMVs negatively correlated with clotting time, regardless of whether they exposed PS (38). In addition to the effects of RMV-associated PS, RMVs also sustainably consume anticoagulant protein C and protein S, contributing to prothrombotic conditions. Furthermore, RMVs derived from aged erythrocytes can induce pulmonary microthrombi through p-selectin in mice (39). Moreover, RMVs were one of the most abundant MVs types in the plasma and atherosclerotic plaques in patients with coronary heart diseases (40). Studies also found that RMVs exert a more pronounced influence on coagulation than PMVs (41, 42) . All of the above studies suggest that RMVs have significantly procoagulant activity via different mechanisms. In our study, we found higher levels of Annexin V^+^RMVs in SLE-PAH, along with increased RDW-SD and RDW-CV, when compared with SLE-non-PAH, suggesting that the importance of RBCs and their released procoagulant RMVs in SLE-PAH development. However, there was no shortening of coagulation time in the SLE-PAH group, and even prolonged PT and PT-INR. This paradoxical finding could be attributed to the fact that SLE-PAH patients were more likely to receive anticoagulant therapy (9 of 18 in SLE-PAH *vs.* 1 of 36 in SLE-non-PAH patients) due to their more severe overall clinical conditions.

It’s known that activation of leukocytes and platelets is enhanced in PAH patients (43–45). Importantly, our study demonstrated increased levels of LMVs and PMVs in SLE-PAH patients. These results are consistent with previous research in PAH (29). Furthermore, PMVs can mediate leukocyte-platelet interaction, triggering activation of leukocytes and subsequent release of LMVs (50). In addition to the procoagulant properties of MV-associated PS (13, 14), previous studies have shown that LMVs and PMVs can induce endothelial injury, release of EMVs and inflammatory factors, proliferation of vascular smooth muscle cells, and intimal thickening (46–49). As circulating EMVs result from activated or apoptotic vascular ECs or damaged vascular endothelium, circulating EMVs could serve as a biomarker of vascular injury, indicating the pathologic conditions with impaired vasculature in patients (34). Many sentinel events, including those triggered by LMVs and PMVs, could induce endothelial injury, increasing the release of EMVs and worsening endothelial dysfunction. Since CD144 is a constituent marker of ECs, highly increased levels of CD144^+^EMVs in circulation indicate impairment of vascular endothelium and their contribution to PAH development in SLE-PAH patients. Furthermore, Amabile reported a positive correlation between blood levels of CD144^+^EMVs and hemodynamic severity of PAH (30). In line, we also found that the elevated levels of EMVs in patients with SLE-PAH were positively correlated with PASP and RVD, suggesting that circulating EMV levels may reflect the severity of SLE-PAH. However, no correlation between EMV levels and RHC parameters was found. Interestingly, our results indicate that there was a close relationship between PMVs, EMVs, and LMVs, and their interactions synergistically participating the SLE-PAH development. There were good correlations between these three subgroups of MVs which was consistent with previous studies (29). All the above indicates that systemic inflammation, endothelial injury, and hypercoagulation state co-exist simultaneously and interact each other during PAH development in SLE-PAH. Although no distinction was found in conventional blood cell counts, such as WBC, RBC, and platelets, between SLE-non-PAH and SLE-PAH patients, the levels of LMVs, RMVs, and PMVs were significantly elevated in SLE-PAH patients, indicating that circulating MVs, rather than their parental cells, may be more indicative of the underlying pathological processes in SLE-PAH. Furthermore, statistical analysis also showed no relationship between the levels of circulating MVs and the numbers of their corresponding parental cells in SLE patients with or without PAH (P >0.05, data not shown).

A recent study found that SLE-PAH could be classified into vasculitic and vasculopathic subtypes based on clinical characteristics in patients (51). SLEDAI>9 was an independent predictor for the high-risk vasculitic subtype SLE-PAH (51). The vasculitic subtype of SLE-PAH patients, with more intensive systemic inflammatory responses and more severe systemic symptoms, are more likely to respond to intensive immunosuppressive therapy, making the PAH process reversible in these patients (52). In contrast, the vasculopathic subtype, however, tends to be non-inflammatory vascular remodeling during its PAH pathogenesis. The above classifications are consistent with the results of the present study. Accordingly, the subgroups were divided according to SLEDAI-2K in our study. When the disease activity was low (SLEDAI-2K<10), levels of circulating EMVs, PMVs, and LMVs in SLE-PAH patients showed no difference compared to SLE patients without PAH, while their Annexin V^+^MVs increased significantly. This suggested that the inflammatory responses were not intensive although their hypercoagulability was more pronounced when SLE was not severe. In addition, their levels of RMVs also increased, suggesting that the role of erythrocytes and the RMVs in PAH pathogenesis under these conditions which deserves more research in the future. In contrast, the levels of LMVs, PMVs, RMVs, Annexin V^+^MVs, and Annexin V^+^RMVs were increased in the SLE patients with SLEDAI-2K≥10. This fact indicates that this group of patients presented as vasculitic PAH with not only hypercoagulation state, but also hyperinflammatory state with more pronounced activation of leukocytes, platelets, and erythrocytes *in vivo* in these patients.

PASP is the most important indicator in UCG for screening and assessment of PAH. In our study, circulating LMVs, RMVs, EMVs, and AV^+^MVs were positively correlated with PASP in all SLE patients, suggesting the potential contributions of these circulating MVs to the development of PAH in SLE patients. Importantly, circulating EMVs were positively correlated with PASP in SLE-PAH patients, suggesting the value of EMVs in assessment of PAH severity in patients with SLE-PAH. As the most intuitive indicator of right ventricular dilation, the increased RVD was considered to be a poor and indirect prognostic marker in PAH patients (53). Our results showed that PMVs and EMVs were positively correlated with RVD in patients with SLE-PAH, indicating that platelet- and endothelial-derived MVs are also valuable in assessing the severity of right heart dysfunction, which may however be the consequences of the increased PASP in these patients.

To further evaluate the usefulness of circulating MVs measured in our study, ROC curve analysis indicated that all single-labeled MVs and dual-labeled Annexin V^+^RMVs are valuable to serve as potential biomarkers to identify PAH in SLE patients. Annexin V^+^ MVs was the best predictor of PAH in patients with SLE for both specificity and sensitivity, indicating the importance of PS-exposing MVs in PAH development. In addition, circulating LMVs were significant predictors of SLE-PAH, suggesting the importance of inflammatory leukocytes and their released LMVs in pathogenesis of SLE-PAH. Furthermore, logistic regression analysis indicated that circulating LMVs, RMVs, and anti-NRNP antibody, as well as serositis were independent predictors of PAH in SLE. Among these variables, anti-nRNP antibody and serositis have been widely recognized as predictors of PAH (54, 55). Most importantly, our work demonstrated that levels of LMVs and RMVs could also be independent biomarkers of SLE-PAH. The ability of MVs to identify PAH in patients with SLE has not yet been previously reported.

This study however has some limitations. First, owing to the fact of rare incidence of PAH in SLE, the number of SLE-PAH patients recruited to this study was limited. However, we strictly matched these subjects with SLE patients without PAH and healthy controls, which could eliminate partial bias to a certain extent. Second, only seven SLE-PAH patients had RHC results at the time of blood sample collection. The criteria for SLE-PAH patients included in this study were RHC compliance or PASP > 50 mmHg, as indicated by UCG. Therefore, 11 patients were not tested for RHC owing to factors such as disease intolerance or patient willingness. However, the UCG results and comprehensive clinical evaluation of these patients were highly indicative of PAH. Owing to the limited numbers of RHC results, we explored the relationship between PAH-associated UCG parameters and MV subgroups in the correlation analysis to compensate for the deficiency of incomplete RHC data to some extent. In addition, studies on the mechanisms of MV in various subgroups, especially RMV, in SLE-PAH are ongoing, and we will conduct follow-up reports.

In conclusion, this study demonstrated that all single-labeled LMVs, PMVs, RMVs, EMVs and Annexin V^+^ MVs, as well as dual-labeled Annexin V^+^ RMVs were significantly increased in SLE-PAH patients compared to SLE-non-PAH patients. In patients with inactive/mild SLE, the procoagulant activity of MVs increased in PAH patients. In patients with moderate/high SLE, both procoagulant activity and hyperinflammatory state mediated by MVs were evident in patients with PAH. Therefore, our study indicated that specific subgroups of MVs are involved in the development of PAH in SLE patients. In addition to the widely reported anti-nRNP antibody and serositis, our study demonstrated that circulating LMVs and RMVs are independent biomarkers of SLE-PAH. This work indicated the clinical usefulness of MVs in identification, diagnosis, and management of PAH in patients with SLE.

### Ethics statement

The studies involving human participants were reviewed and approved by the Ethics Committee of Tianjin Medical University General Hospital, and all patient/participants provided their written informed consent in accordance with the Declaration of Helsinki.

## Data Availability

All data produced in the present study are available upon reasonable request to the authors

## Author contributions

Zhe Ding: Methodology, Validation, Formal analysis, Resources, Writing - Original Draft. Fumin Qi: Formal analysis, Investigation, Data Curation. Li Liu: Methodology, Resources. Na Zhang: Conceptualization, Investigation. Xing Lyu: Methodology, Data Curation. Wenwen Sun: Methodology, Data Curation. Jun Du: Investigation. Haoming Song: Resources. Hou Hou: Investigation, Resources. Ying Guo: Investigation. Xiaomei Wang: Investigation. Minglin Liu: Conceptualization, Supervision, Writing - Review & Editing. Wei Wei: Conceptualization, Supervision, Project administration.

## Acknowledgement

Funding: This work was supported by the Tianjin Municipal Health Commission Funding grant (grant numbers KJ20028), National Natural Science Foundation of China (grant numbers 81370422), and Lupus Research Alliance (grant numbers 416805).

